# Comparative Efficacy and Safety of Pharmacological Managements for Hospitalized COVID-19 Patients: Protocol for Systematic Review and Trade-Off Network Meta-Analysis

**DOI:** 10.1101/2020.05.18.20103697

**Authors:** Min Seo Kim, Won Jun Kim, Min Ho An

## Abstract

Coronavirus-Disease 2019 (COVID-19) is the clinical disease caused by the SARS-CoV-2 virus, the infectious agent causing the ongoing pandemic that has impacted the lives of hundreds of millions of people in almost every nation worldwide. It is a potentially fatal disease to many vulnerable patients including the elderly and those with chronic illnesses; but because this virus is a novel one, there are no firmly established treatment protocols. Many treatment methods are being investigated worldwide, and scientific conclusions drawn from these endeavors are crucial for healthcare professionals in combating this disease. In this network meta-analysis, we focus specifically on the pharmacologic agents that have been investigated for the treatment of COVID-19 and aim to produce a comprehensive picture of the evidence from current data in order to produce relevant insights on the comparative efficacy and safety profiles of various pharmacologic agents against COVID-19.

## INTRODUCTION

Coronavirus Disease 2019 (COVID-19) is the disease caused by the virus SARS-CoV-2 first discovered in Wuhan, China in 2019. It has caused a global pandemic affecting almost everyone in the world and causing millions of confirmed cases and hundreds of thousands of deaths (1). While many infected patients experience self-limiting course of the disease, many – especially the elderly and those with underlying illnesses – require hospitalization and intensive care as the virus causes decompensation of the body’s systems causing potentially lethal complications such as acute respiratory distress syndrome (ARDS). Therefore, treatments to reduce death and decompensation while expediting viral clearance and clinical recovery are crucial. However, as of May 16^th^, 2020 while many pharmaceutical agents have been and are being investigated worldwide for efficacy and safety in COVID-19 patients, none have been shown to have clear superiority over others. Therefore, in this network meta-analysis (NMA), we aim to compare the multiple treatment options that are currently being investigated for COVID-19 with the goal of synthesizing the scattered research results on the pharmacological treatment of COVID-19, determining which agents have demonstrated efficacy, and finally comparing agents against each other to determine which ones are most effective and safe for hospitalized COVID-19 patients based on data accrued so far.

## METHODS

We will conduct this NMA based on the Preferred Reporting Items for Systematic Reviews and Meta-Analyses (PRISMA)-NMA guidelines (2). This investigation is currently under review for registration on the International Prospective Register of Systematic Reviews (PROSPERO). If deviations are made to the prospectively registered protocol, they will be reported in the final publication.

### Eligibility Criteria

#### Types of Participants

Participants included in our analysis will be those with confirmed infection with SARS-CoV-2 and hospitalized for the management of the resultant disease COVID-19. Patients of COVID-19 with mild symptoms do not require hospitalization are not eligible for our analysis.

#### Types of Interventions

Our analysis will include all pharmacological treatments that have been investigated for COVID-19: antivirals, immunomodulators, antibiotics, antibodies, corticosteroids, antimalarials, etc. Our inclusion criteria will not have restrictions on dosage, regimen, dosing interval, route of administration, or intervention duration.

#### Types of Studies

We will include randomized controlled trials (RCTs), non-randomized controlled trials, prospective cohort studies, retrospective cohort studies, and preprints and unpublished studies. For preprints and unpublished studies, the authors of each study will be contacted to identify any critical changes made to the studies after the visualization of the preprint. Following studies will be excluded: pilot studies with small sample sizes, literature review of qualitative syntheses and theoretical explanations, opinions or comments, animal or in vitro experiments.

### Primary Outcomes

The main outcomes that will be assessed are mortality rate, intubation rate, and total adverse event rate for each pharmaceutical agent. Odds ratios (ORs) and relative risk (RR) will be calculated as appropriate.

### Secondary Outcomes

Secondary outcomes that will be assessed are time to viral clearance (demonstrated by negative conversion on real-time polymerase chain reaction [RT-PCR]), duration of hospital stay, duration of intensive care unit (ICU) stay, duration of ventilation, rate of indication for renal-replacement therapy, rate of indication for non-invasive ventilator support, and clinical improvement. Data on cardiac arrest, arrhythmia, and QTc prolongation will also be collected for safety evaluation of treatments. OR, RR, and mean difference (MD) will be used to measure effect size.

### Search Methods for Identification of Studies

#### Electronic Database Search

We will search PubMed, Google Scholar, MEDLINE, the Cochrane Library (Cochrane database of systematic reviews), CINAHL, Scopus, Embase from inception to May 2020. For unpublished studies and registered ongoing trials, we will also search medRxiv, bioRxiv, arXiv, International Clinical Trials Registry Platform (http://apps.who.int/trialsearch), Epistemonikos COVID-19 LOVE platform (http://app.iloveevidence.com/loves/), and ClinicalTrails.gov (http://clinicaltrials.gov/). We will use appropriate MeSH terms as exemplified in the following example:

(covid[ti] OR SARS-CoV-2[ti] OR corona*[ti]) AND (treat*[ti] OR EIDD[tiab] OR antiviral[tiab] OR flu[ti] OR favipiravir[tiab] OR avigan[tiab] OR umifenovir[tiab] OR arbidol[tiab] OR chloroquine[tiab] OR hydroxychloroquine[tiab] OR azithromycin[tiab] OR antibiotics[tiab] OR ebola[ti] OR remdesivir[tiab] OR HIV[ti] OR kaletra[tiab] OR lopinavir[tiab] OR ritonavir[tiab] OR actemra[tiab] OR tocilizumab[tiab] OR interleukin[tiab] OR sarilumab[tiab] OR kevzara[tiab] OR corticosteroid[tiab] OR prednisolone[tiab] OR ACEi[tiab] OR angiotensin[tiab] OR malari*[tiab]) AND (label*[tiab] OR retrospective[tiab] OR randomi*[tiab] OR observational[tiab])

The authors will also manually search reference lists of review papers and meta-analyses.

The search will be conducted independently by three authors (MK, MA, WK) and discrepancies resolved by discussion.

### Data Collection and Analysis

#### Study Selection

Titles, abstracts, and keywords of each study will be reviewed independently by three authors (MK, MA, WK) for full-text screening. Full-text screening will also be conducted by each of the three authors independently for final inclusion in the analysis. Discrepancies will be resolved through discussion.

#### Data Collection

Data extraction will also be conducted independently by the three authors using a predetermined data extraction spreadsheet. Discrepancies will be resolved through discussion.

#### Risk of Bias

Two authors (MA, WK) will independently assess risk of bias (RoB) in included studies using the Cochrane Collaboration’s tool for RoB assessment (3). Any discrepancies will be resolved through discussion and/or arbitration by a third author (MK). For RCTs, the following domains, as described in the Cochrane Handbook for Systematic Reviews of Interventions, will be assessed for each article to determine RoB: 1) random sequence generation, 2) allocation concealment, 3) blinding of participants and personnel, 4) blinding of outcome assessment, 5) incomplete outcome data, 6) selective reporting, and 7) other biases. For observational studies, RoB will be assessed on the same domains except random sequence generation and allocation concealment as these are not applicable to observation studies.

#### Study Quality Assessment

Newcastle-Ottawa scale (NOS) and Jadad quality scale will be used for study quality assessment of observational studies and RCTs, respectively.

#### Strength of Evidence

The strength of evidence for each main/primary outcome will be evaluated with the Agency for Healthcare Research and Quality (AHRQ) approach and will be classified as high, moderate, low or insufficient. Strength of evidence (SoE) evaluates potential weaknesses that must be taken into account when interpreting the results including small sample size effect, unrealistically large or small odds ratios with extended 95% confidential intervals, high risk of bias of individuals studies composing individual outcomes, inconsistency between direct and indirect evidence, and several other reporting biases.

### Data Items

#### Bibliometric Data

Authors, study design, digital object identifier (DOI), region, publication journal, funding sources, conflicts of interest.

#### Methodological Data

Study design, randomization and allocation methods, duration of enrollment and trial

#### Outcome Data

Baseline characteristics of included patients including severity classification, number of patients in each treatment arm, endpoints (i.e. the primary and secondary outcomes – mortality rate, intubation rate, adverse event rate, time to viral clearance, etc.), details of pharmacologic intervention (dose, duration, compound, regimen, route of administration, etc.).

### Statistical Analysis

#### Network Meta-Analysis

We will conduct a random-effects NMA within a frequentist framework using STATA (Stata Corp, College Station, TX, US, version 15.0) and R (version 3.6.0) software. We will duplicate the results by analyzing an identical data set independently in the two different software packages and cross-check whether the results are comparable to minimize error. Indirect and mixed comparison will be accomplished through the mvmeta command and self-programmed routines of STATA, and the netmeta package of R. Estimated pooled effect sizes from the network meta-analysis will be presented as MD or standardized mean difference (SMD) for continuous outcomes and ORs for dichotomous outcomes with 95% CIs. When a continuous variable is presented as median (interquartile range), it will be converted to a mean (standard deviation) (4, 5).

#### Treatment Ranking

We will use SUCRA scores to provide an estimate as to the ranking of treatments.

#### Heterogeneity Assessment

Statistical heterogeneity will be estimated using Higgins I^2^ and the Cochran Q statistic. We will use the restricted maximum likelihood method to evaluate heterogeneity, assuming a common heterogeneity variable for all comparisons (the tau value) and compute I^2^ and its 95% CI.

#### Inconsistency

Global inconsistencies between direct and indirect comparisons will be evaluated using a design-by-treatment model (6). Local inconsistencies will be assessed by a loop-specific approach for every closed triangular or quadratic loop and by a node-splitting method. The net heat plot will be constructed using the netmeta package of R (version 3.5.1) to visualize the inconsistency matrix and detect specific comparisons which contribute to large inconsistencies(7).

#### Publication Bias

A comparison-adjusted funnel plot will be constructed to assess publication bias.

#### Subgroup and Sensitivity Analyses

Subgroup analyses are planned for the following subgroup classifications, if applicable:

severity of disease e.g. moderate versus severe/critically ill
study design e.g. RCTs plus large cohort observational studies (>100 or >200 cases for each arm) versus RCT plus all observational studies
publication status e.g. published studies only versus published plus unpublished studies (i.e. pre-print).

If subgroup analyses are not applicable, sensitivity analyses will be conducted for 1) RCT plus large cohort observational studies (>100 or > 200 cases for each arm) only, and 2) published studies only.

Some of these subgroup and sensitivity analyses will be placed in supplementary materials.

## DISCUSSION

COVID-19 is an ongoing global pandemic, and scientific conclusions about potential treatment approaches to the disease is a time-sensitive issue that has the potential to impact the rapidly increasing number of infected patients and their healthcare systems around the world. This analysis focuses on the pharmacologic interventions that have been proposed as potential treatment agents for COVID-19 and aim to compare their efficacy and safety through a network meta-analysis.

## Data Availability

Anyone can share this material, provided it remains unaltered in any way, this is not done for commercial purposes, and the original authors are credited and cited.

## ACKNOWLEDGEMENTS

None.

## AUTHOR STATEMENT

None

## AUTHOR APPROVAL

All authors have been seen and approved the manuscript

